# National Consumption of Antimalarial Drugs and COVID-19 Deaths Dynamics: An Ecological Study

**DOI:** 10.1101/2020.04.18.20063875

**Authors:** Maxime Izoulet

## Abstract

COVID-19 (Coronavirus Disease-2019) is an international public health problem with a high rate of severe clinical cases. Several treatments are currently being tested worldwide. This paper focuses on anti-malarial drugs such as chloroquine or hydroxychloroquine. We compare the dynamics of COVID-19 daily deaths in countries using anti-malaria drugs as a treatment from the start of the epidemic versus countries that do not, the day of the 3rd death and the following 10 days. We then use a ARIMA modeling to realize a short-term forecast of deaths dynamics for each group. We show that the first group have a much slower dynamic in daily deaths that the second group. This ecological study is of course only one additional piece of evidence in the debate regarding the efficiency of anti-malaria drugs, and it is also limited as the two groups certainly have other systemic differences in the way they responded to the pandemic, in the way they report death or in their population that better explain differences in dynamics. Nevertheless, the difference in dynamics of daily deaths is so striking that we believe it is useful to present these results as a clue in the researches about the efficiency of hydroxychloroquine. In the end, this data might ultimately be either a piece of evidence in favor or anti-malaria drugs or a stepping stone in understanding further what other ecological aspects place a role in the dynamics of COVID-19 deaths.

## 1. Introduction

COVID-19 (Coronavirus Disease-2019) is an international public health problem with a high rate of severe clinical cases. Several treatments are currently being tested worldwide. This paper focuses on anti-malarial drugs such as chloroquine or hydroxychloroquine, which have been currently reviewed by a systematic study as a good potential candidate^1^ and that has been reported as the most used treatment by a recent survey of physicians^2^. We compare the dynamics of COVID-19 daily deaths in countries using anti-malaria drugs as a treatment from the start of the epidemic versus countries that do not, the day of the 3rd death and the following 10 days. We show that the first group have a much slower dynamic in daily deaths that the second group. This ecological study is of course only one additional piece of evidence in the debate regarding the efficiency of anti-malaria drugs, and it is also limited as the two groups certainly have other systemic differences in the way they responded to the pandemic, in the way they report death or in their population that better explain differences in dynamics (systematic differences that may also explain their choice to rely on anti-malaria drugs in the first place). Nevertheless, the difference in dynamics of daily deaths is so striking that we believe that the urgency context commands presenting the results before delving into further analysis. In the end, this data might ultimately be either a piece of evidence in favor or anti-malaria drugs or a stepping stone in understanding further what other ecological aspects place a role in the dynamics of COVID-19 deaths.

## 2. Method

In this study, we set up two groups of 16 countries and study the dynamics of the number of deaths between the day of the 3rd death and the following 10 days. The first group is made up of countries that we know use or produce chloroquine or hydroxychloroquine on a massive scale during this period. The second group consists of countries that did not use or produce chloroquine or hydroxychloroquine in large quantities during the period under consideration. When we calculate the averages of each of the two groups, we find very marked differences in their temporal dynamics (see results).

We then use Box and Jenkins’ methodology to apply ARIMA (Auto Regressive Integrated Moving Average) models to these time series, compare the model parameters obtained for each group of countries, and make forecasts of the means of the two groups from these results. Unsurprisingly, the ARIMA models predict a stabilization of the number of deaths for the group of countries using chloroquine and a large increase for the group of countries not using it. The 60 countries most affected by the epidemic (in terms of number of cases) were studied one by one in descending order to determine whether or not they were conducting a national strategy for the large-scale use or production of chloroquine at the beginning of the epidemic in the country (around the 3rd death)^3^. If there was no evidence of such a strategy, or even if sources indicated a strategy to the contrary, the country was classified in the “control group” group, until a panel of 16 countries was obtained in order to have a large sample, provided that daily death data were available for the 10 days following the third death. The second group was constituted with the 16 countries among the 60 most affected in terms of number of cases for which sources indicate the massive use or production of chloroquine at the beginning of the epidemic in the country (around the 3rd death), provided that they have daily death data for the 10 days following the 3rd death. The different groups of countries were constituted according to the information available in the international press on their use or mass production of such drugs over the period under consideration. 16 countries thus constitute each of the two groups (see figure 1 and figure 2).For each of the two groups, the number of daily deaths is noted each day from the 3rd death in the country and the following 10 days. Then the average of the daily deaths is established for each day for each group of countries. For the group without chloroquine, an average is also calculated by removing China and another by removing China and Spain, as these two countries have the two most explosive time series and may be seen as outliers. The trends do not change substantially.

**Figure 1:**
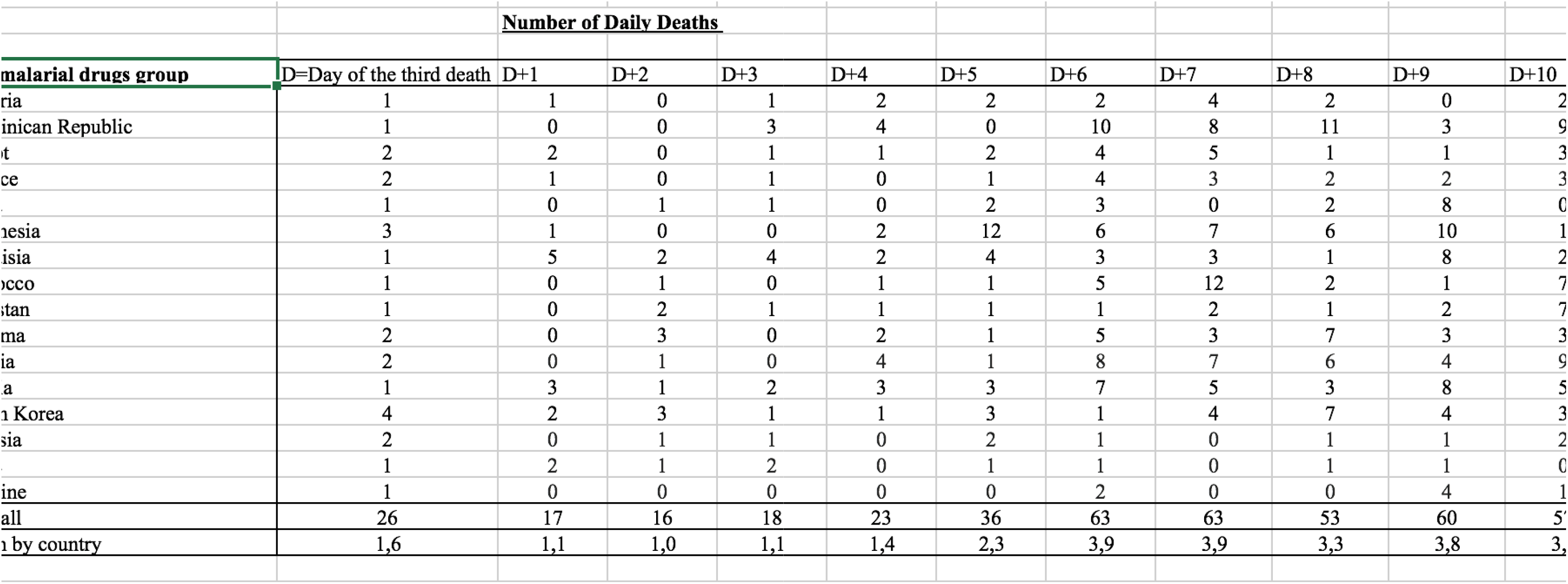
Number of daily deaths after day with 3 deaths, “antimalarial drugs group”

**Figure 2:**
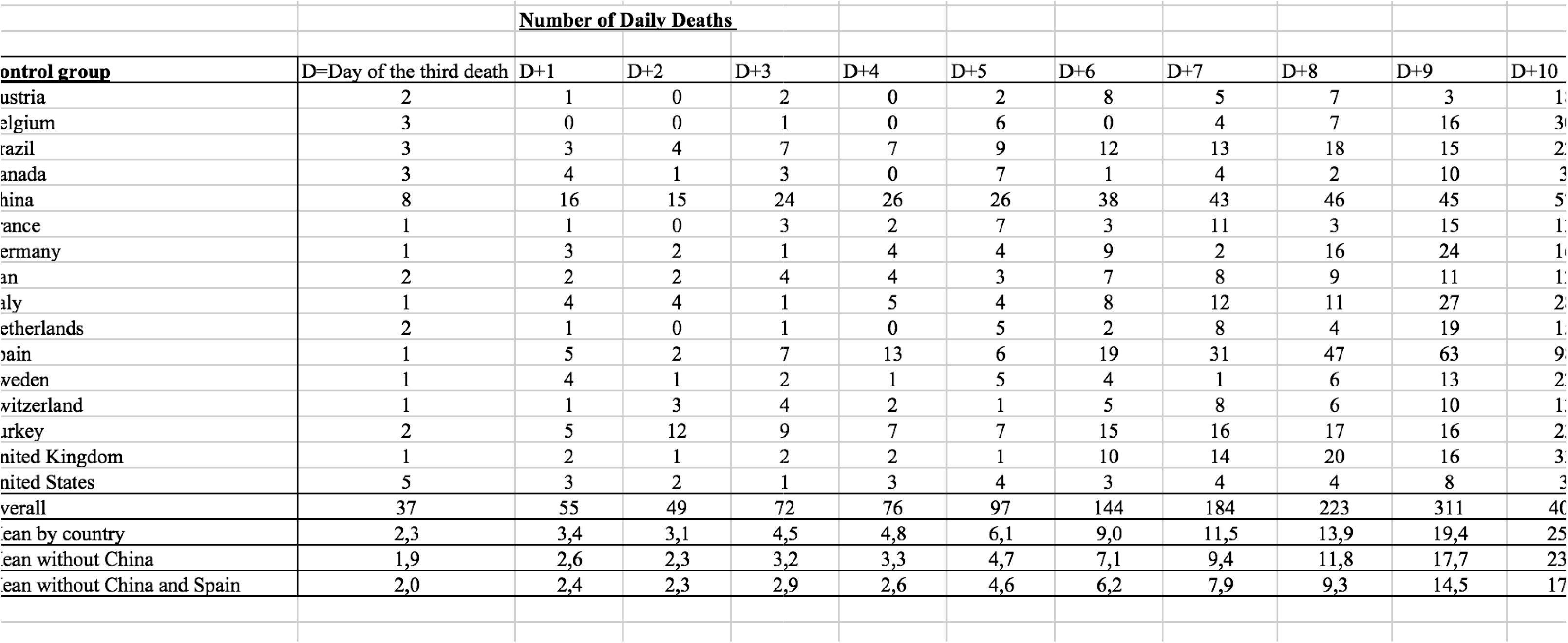
Number of daily deaths after day with 3 deaths, “control group”

## 3. Results

The graphical projection of the mean curves indicates a divergence in the dynamics of the daily death curves of the two groups of countries which is very clear for the period studied (i.e. from the beginning of the epidemic) (**see figure 3**).

**Figure 3:**
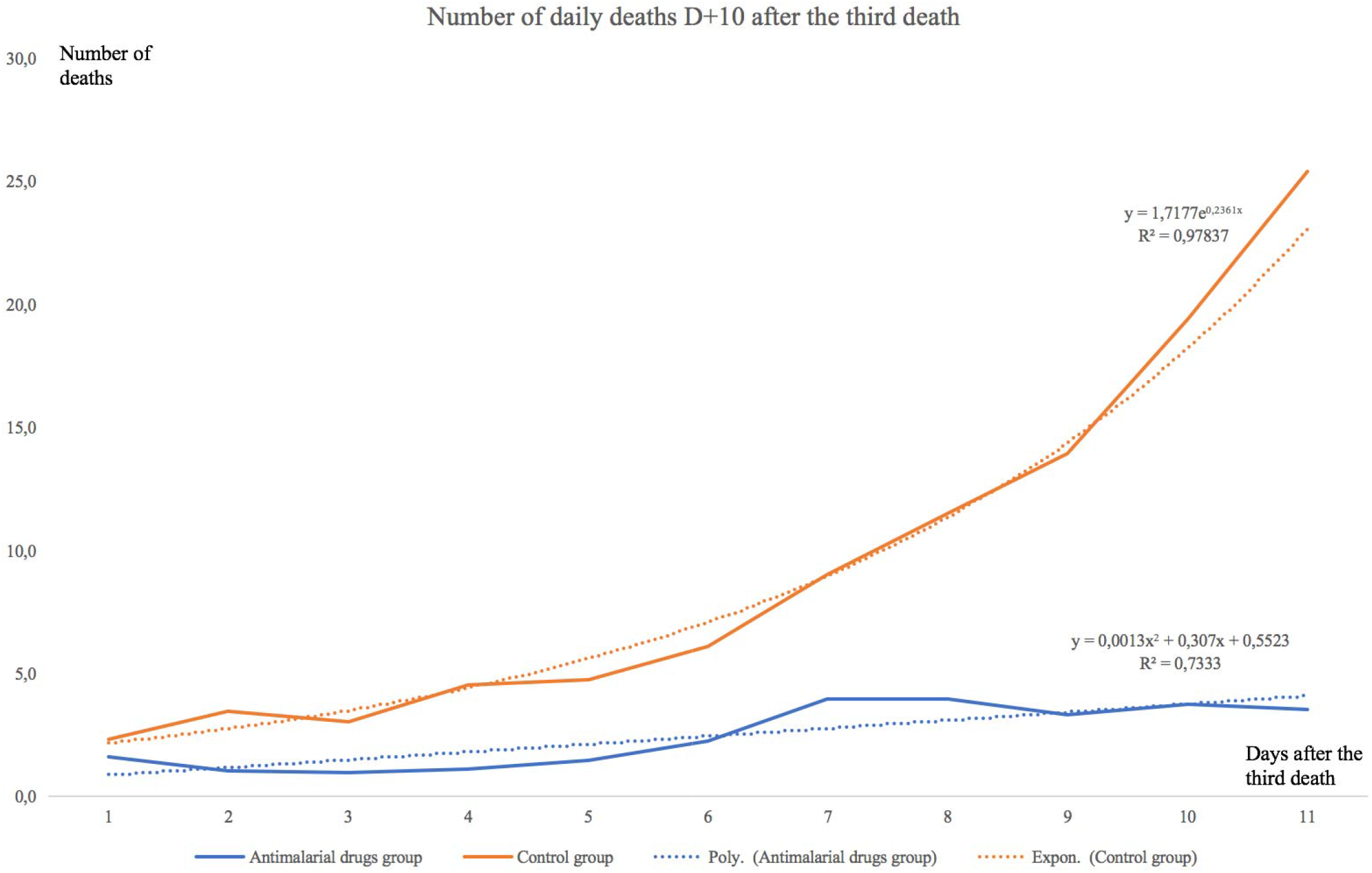
Means of the number of daily deaths for each group

The average curve for countries using antimalarial drugs is rather stable or slightly increasing, the curve for countries not using those treatments is on the contrary strongly increasing. Moreover, the simple regression curves clearly indicate this difference in trend. The average of countries with widespread chloroquine use is fairly well modelled *(R*^2^ = 0,73) by a slightly ascending polynomial regression, whereas the average of countries without chloroquine is very well modelled (*R*^2^ = 0,98) by an exponential regression. Modelling and forecasting using ARIMA (Auto Regressive Integrated Moving Average) models are widely used in time series econometrics. Introduced by Box and Jenkins, they allow an excellent modelling of time series based on the data themselves and without including any theoretical *a priori* on these data. They therefore allow excellent modelling of the internal dynamics of these data and are highly predictive, which tends to validate their relevance. They are widely used in macroeconomics and finance, but also in many other fields, in biology, geophysics, astronomy, etc…

Let’s say an ARIMA (*p,d,q*) process:

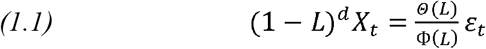

With Ø*_i_*(*i* = 1,…,*p*) the reals corresponding to the autoregressive coefficients, *θ_j_*(*j* = 1,…,*p*) the reals corresponding to the moving average coefficients, of the order of integration *d* and (*ε_t_*~*WN*(0, *σ*^2^)) the residuals behaving as white noise, with zero mean and variance *σ*^2^, constant and less than infinity.

Following Box and Jenkins’ methodology for specifying, estimating and validating the ARIMA modelling, the application to the mean time series of the two groups of countries using the R^4^ software gives the results of **the figure 4**.

**Figure 4:**
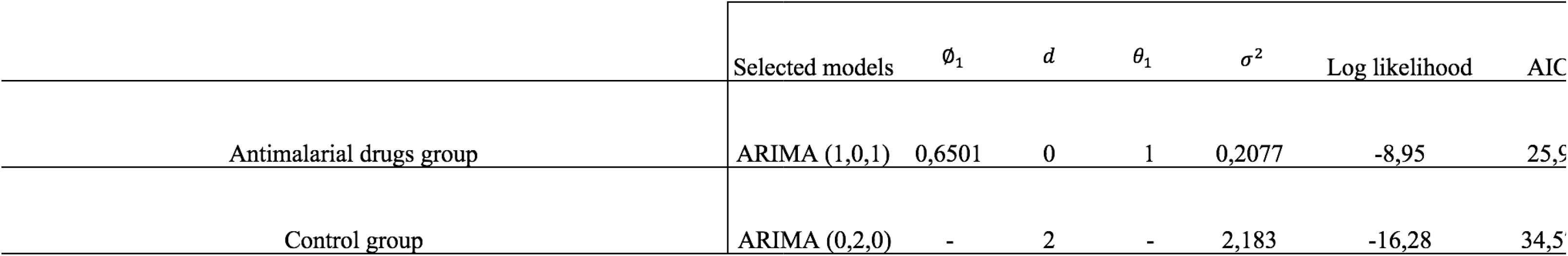
ARIMA parameters specifications and estimations for each group

The Akaike Information Criterion (AIC), for each model selected, is the best relative to other alternative models that were also calculated in this study, i.e. it is closest to zero, indicating the quality of the model specification. This criterion is calculated as follows:

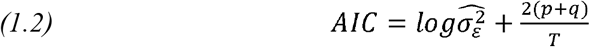

This modelling then allows a 10-day forecast of the evolution of the death dynamics for each of the two groups of countries. We obtain the results in *R* (the first column shows the number of days after the first day with 3 deaths, the second column shows the estimated forecast values, the third and fourth columns show the low and high values of the 80% confidence intervals of the forecast, and the fifth and sixth columns show the 95% confidence intervals)

-For the “antimalarial drugs group”, **see figure 5 and figure 6**.

-For the group “without chloroquine”, **see figure 7 and figure 8**.

**Figure 5:**
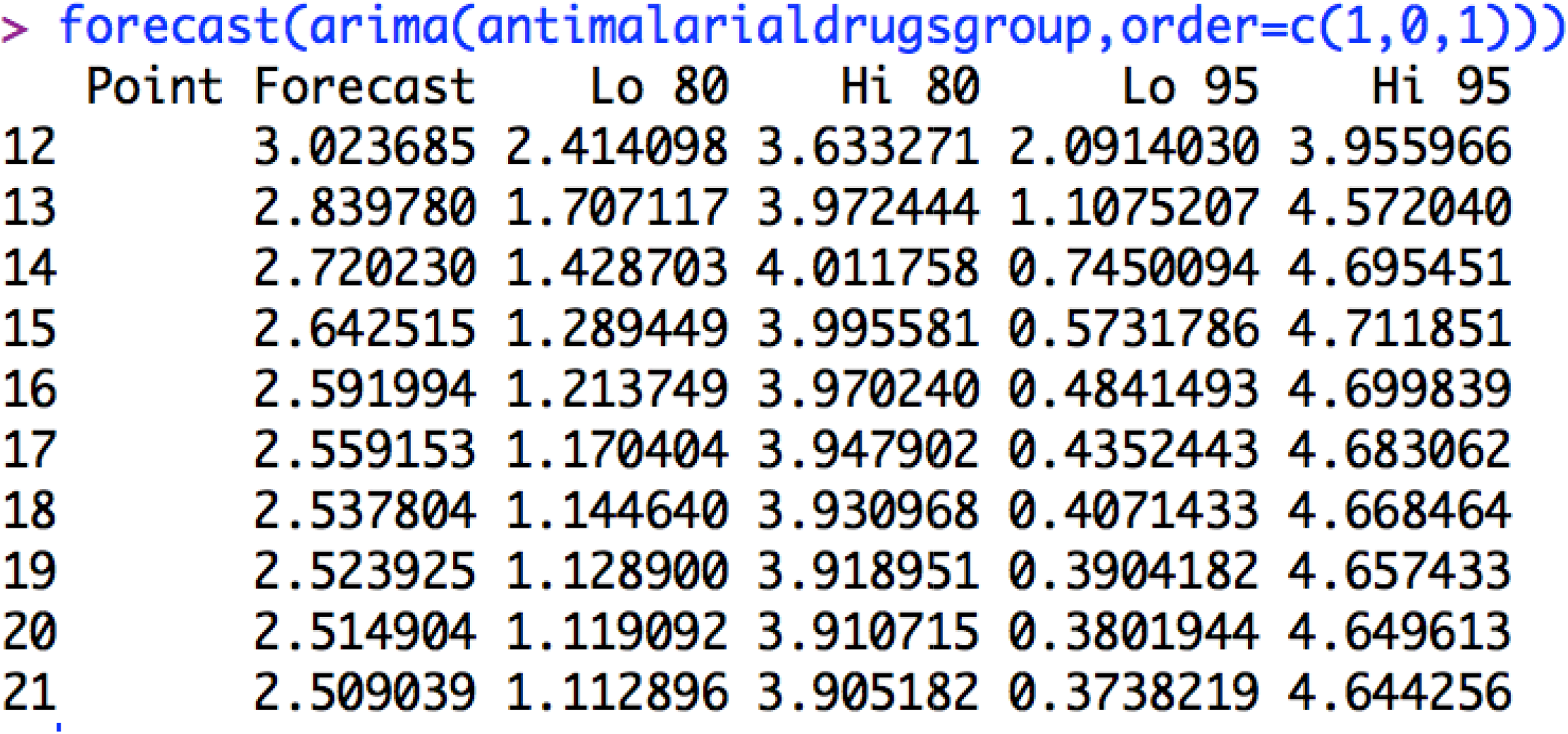
Forecasting values and confidence intervals for an ARIMA (1,0,1) process applied to “antimalarial drugs group”

**Figure 6:**
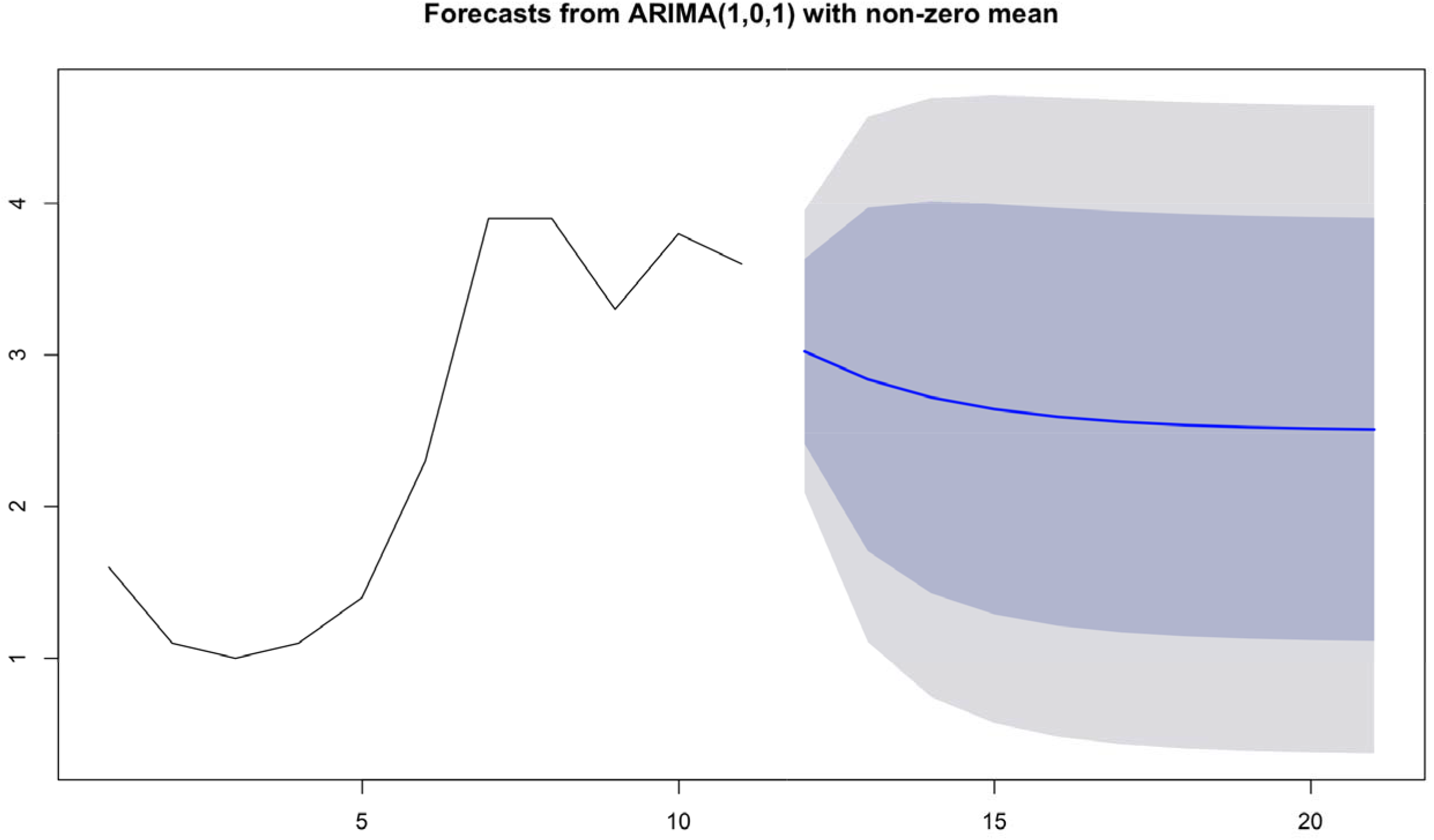
Forecasts plot for “antimalarial drugs group”

**Figure 7:**
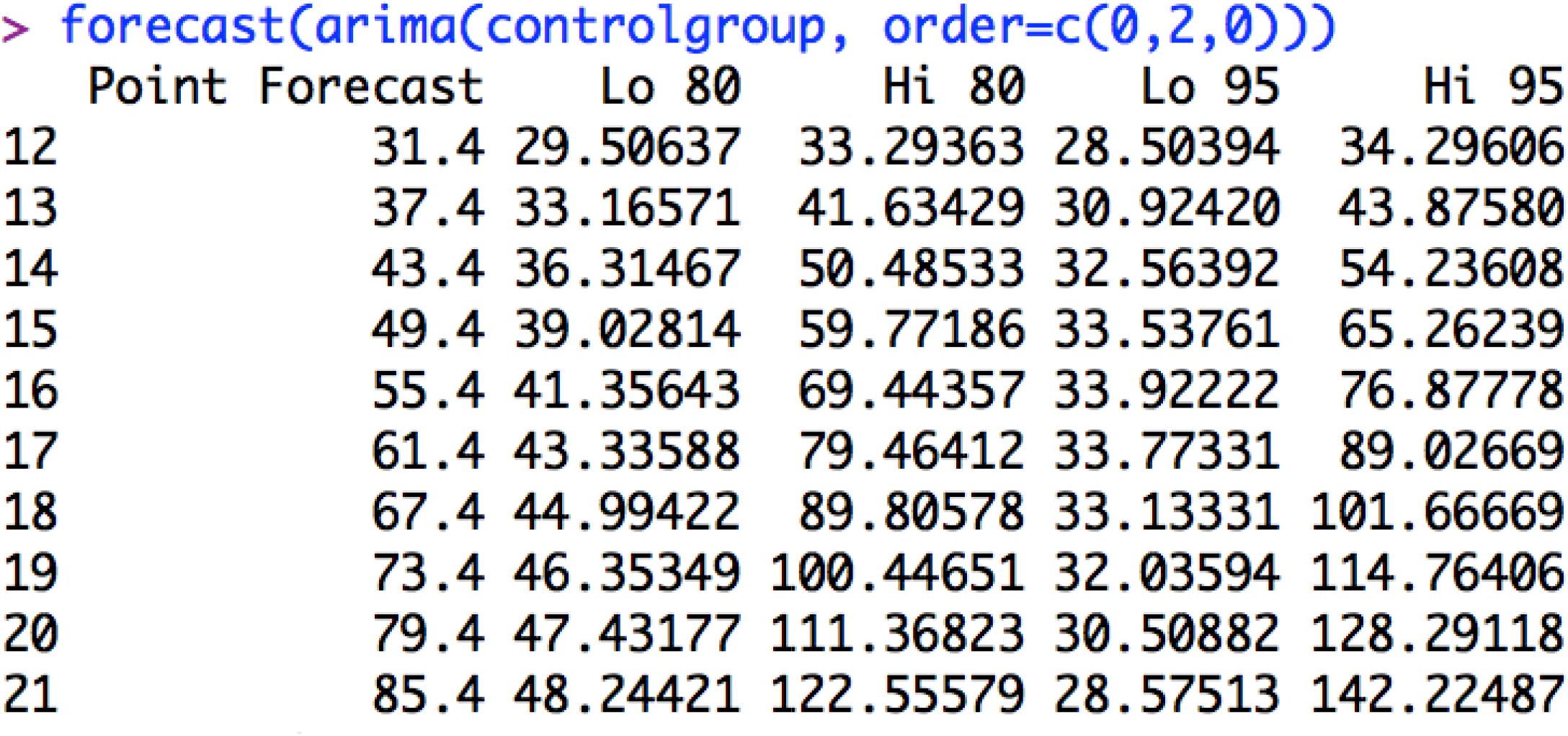
Forecasting values and confidence intervals for an ARIMA (0,2,0) process applied to “control group”

**Figure 8:**
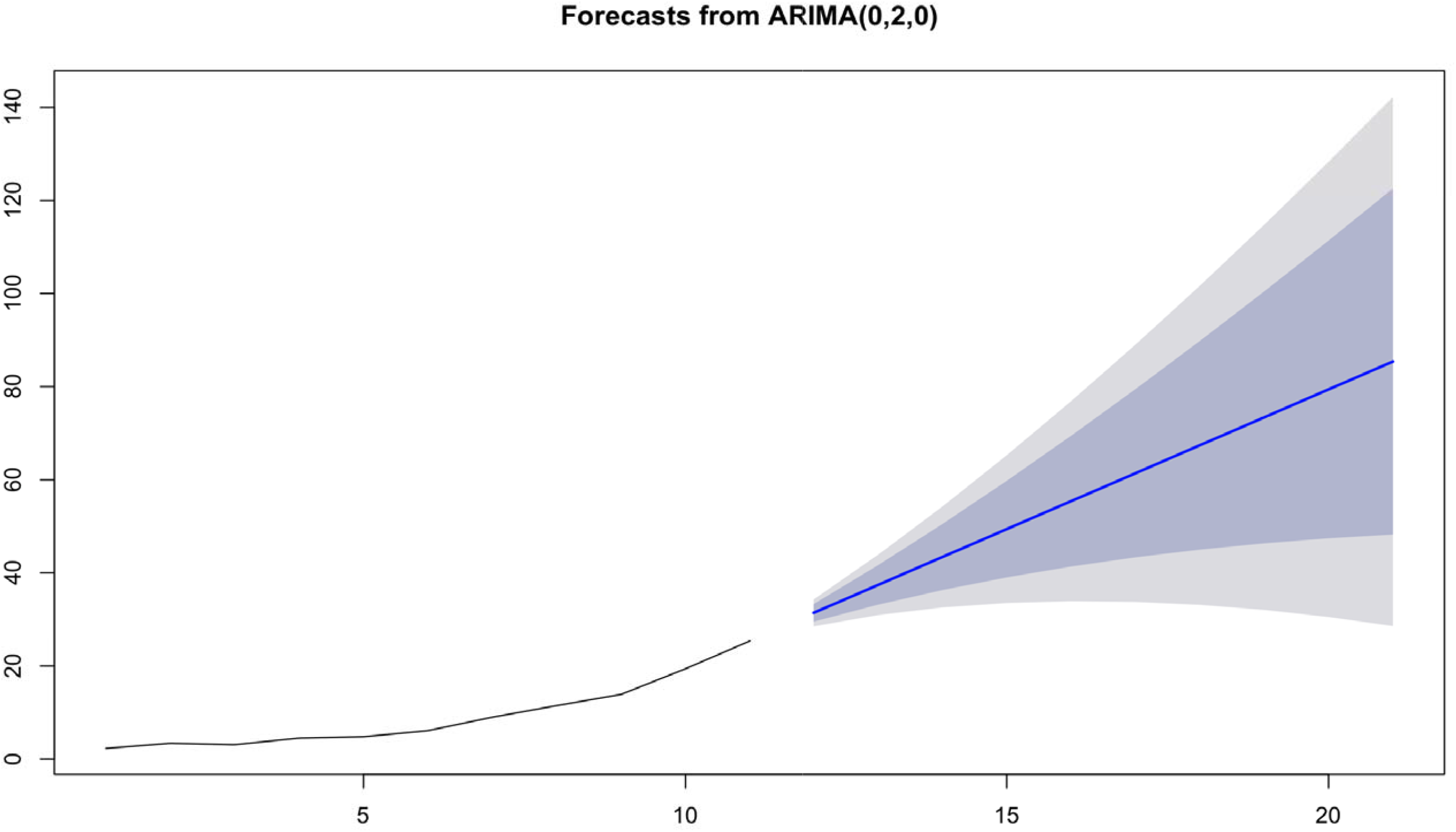
Forecasts plot for “control group”

Forecasts reinforce early visual observations. For the group of countries “antimalarial drugs group”, the forecast of the ARIMA model (1,0,1) indicates a stabilization of the death curve. For the “control group” countries, the ARIMA model’s forecast (0,2,0) indicates a very significant acceleration in the number of deaths. It should be noted that beyond D+10, such an acceleration is already visible in the actual data of many countries for which this statistic is available.

To validate model’s specification, residuals distribution is then tested, in order to control they behave as a white noise, i.e. they are not autocorrelated. This verification is done through the autocorrelations of residuals plotting in *R*.

Autocorrelation function is a *X_t_* process of *k* order that can be writing as follow:

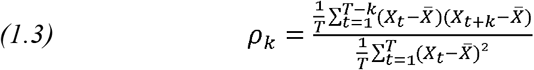

For ARIMA (1,0,1) applied to “antimalarial drugs group”, we obtain autocorrelations of **the figure 9**.

**Figure 9:**
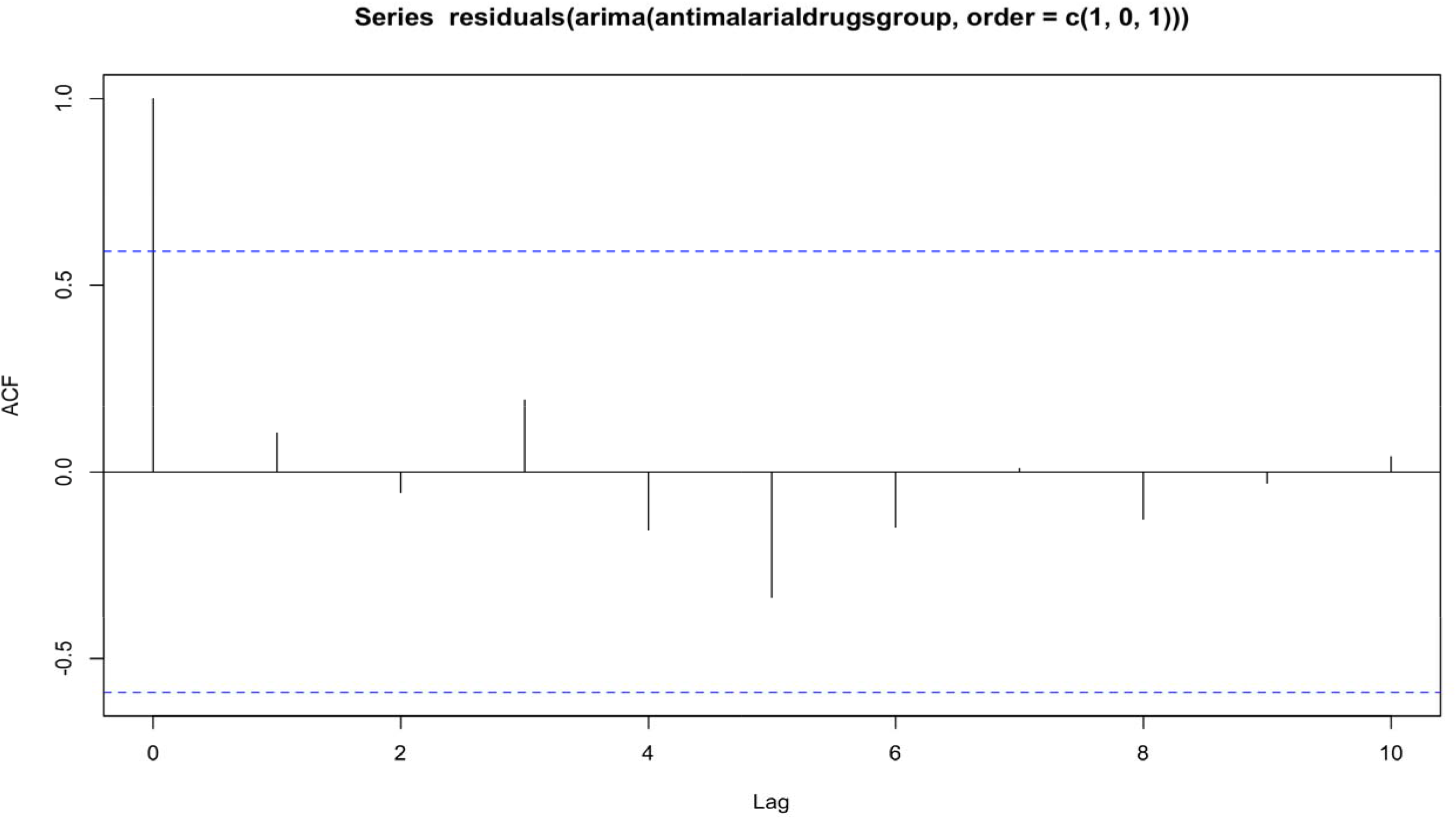
Autocorrelations of residuals for ARIMA (1,0,1) applied to “antimalarial drugs group”

No autocorrelation is significant, residuals are behaving as a white noise, it indicates the validity of the model.

For ARIMA (0,2,0) applied to “control group”, we obtain autocorrelations of **the figure 10**.

**Figure 10:**
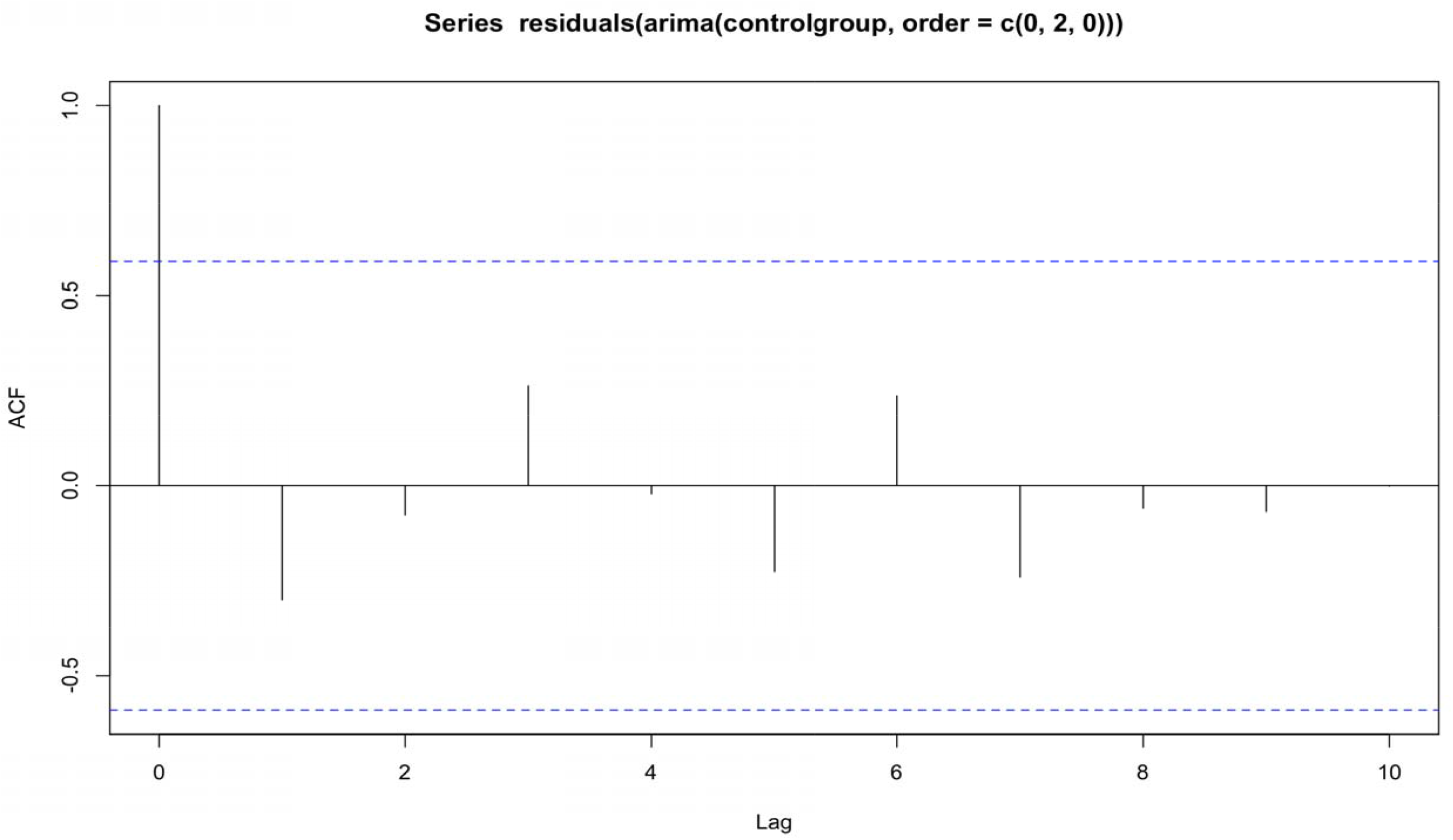
Autocorrelations of residuals for ARIMA (0,2,0) applied to “control group”,

In the same way, no residuals autocorrelation is significant. Residuals are behaving as a white noise, model specification and estimation is then validated.

## 4. Limitations

Introduced in the 1970s by Box and Jenkins^5^, ARIMA models are so-called a-theoretical models^6^, which seek predictive efficiency by focusing on the past data of a time series, without worrying about the causes of these past data. They are therefore not able to explain all the explanatory variables of a temporal evolution, but they are very effective in describing the internal dynamics of the evolution. Nor are they an instrument of proof, but rather a statistical index updating a dynamic. Here they make it possible to highlight two very distinct dynamics from the very first days of the outbreak, which is very useful since this highly contagious epidemic has a strong internal dynamic. They have been already used for modelling the spread of the epidemic, notably in India.^7^

Of course, they do not model, and do not claim to model, all the parameters that explain a temporal evolution. On the other hand, they are often highly predictive^8^ and outweigh many models with more explanatory variables, which is a very important criterion of overall model validity. It should also be noted that while many sources exist to determine the health action of governments, including their use or mass production of chloroquine from the onset of the crisis, quantitative data are lacking and do not allow for more in-depth temporal analyses and causality tests. There also might be systematic differences between the two groups - in particular political differences, urban differences or differences in other strategy aspects such as testing. There is strong evidence for places like South Korea and Japan that mass testing is an effective strategy to control the epidemic, and our study might be a proxy for testing strategies. All these aspects should be examined in a late study.

## 5. Conclusion

We find major differences in death rates, with countries using antimalarial drugs faring better than those which do not. This analysis is of course only one additional piece of evidence in the debate regarding the efficiency of anti-malaria drugs, and it is also limited as the two groups certainly have other systemic differences in the way they responded to the pandemic. Nevertheless, the differences in dynamics is so striking that we believe that the urgency context commands presenting this ecological study before delving into further analysis. In the end, this data might ultimately be either a piece of evidence in favor or anti-malaria drugs or a stepping stone in understanding further what other ecological aspects place a role in the dynamics of COVID-19 deaths.

## Data Availability

All the available data referred in the manuscript are in the data availability link below.

https://www.data.gouv.fr/fr/datasets/daily-deaths-due-to-covid19-by-countries-classed-by-their-using-or-not-of-antimalaria-drugs/#_

## Acknowledgement

I would like to thank all the colleagues of my laboratory (CEMI-EHESS, Paris) for their support and help. I’m also grateful to *Institut Européen d’Etudes du Développement* (IEED) in Paris.

## Statement of Ethics

This study has been conducted ethically, in accordance with the World Medical Association Declaration of Helsinki.

## Author Contributions

Maxime Izoulet has realized the integrality of this paper.

## Potential international reviewers

No potential reviewers have been selected for this article.

### Annexes

**Annex 1:**
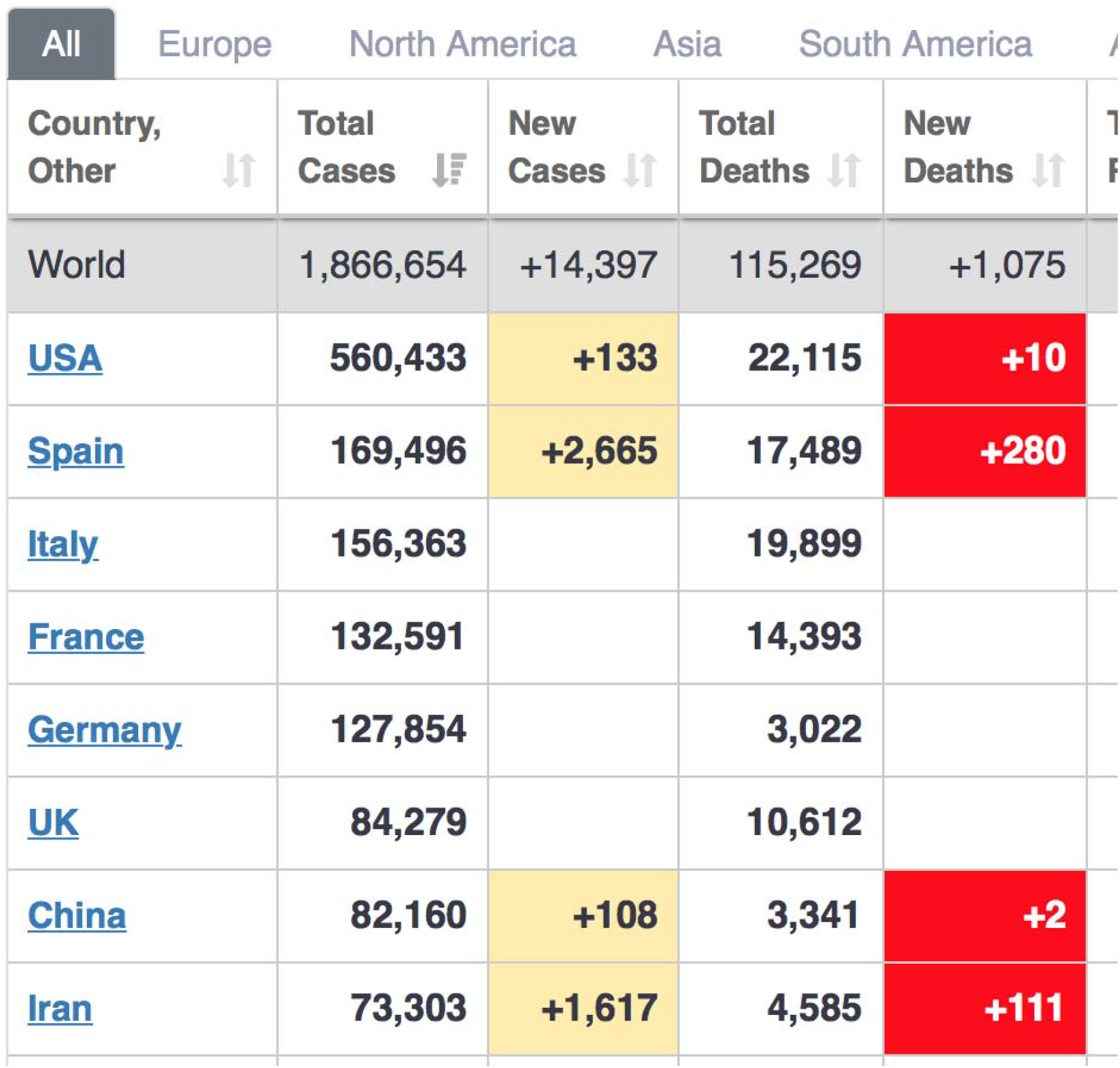
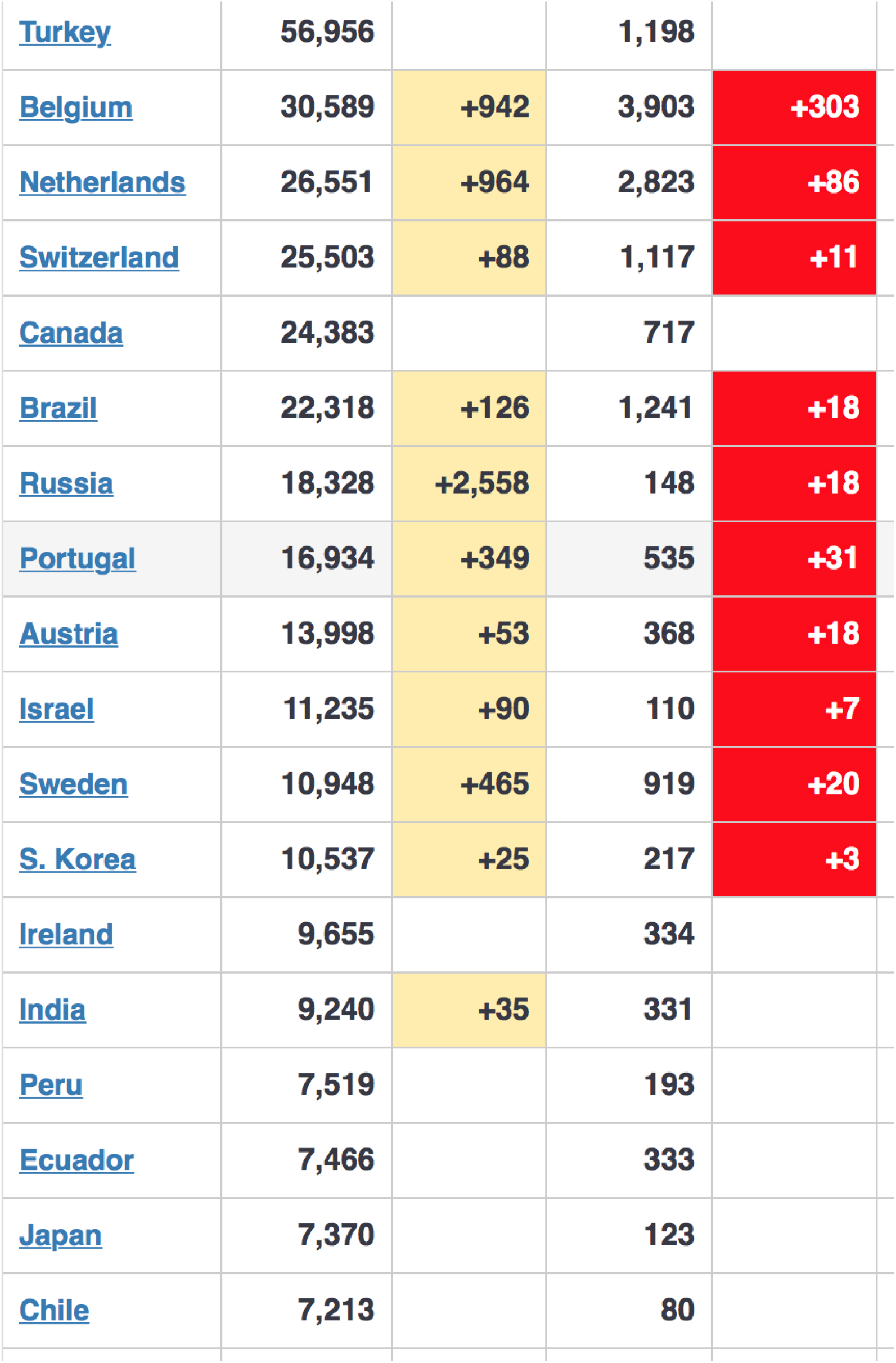
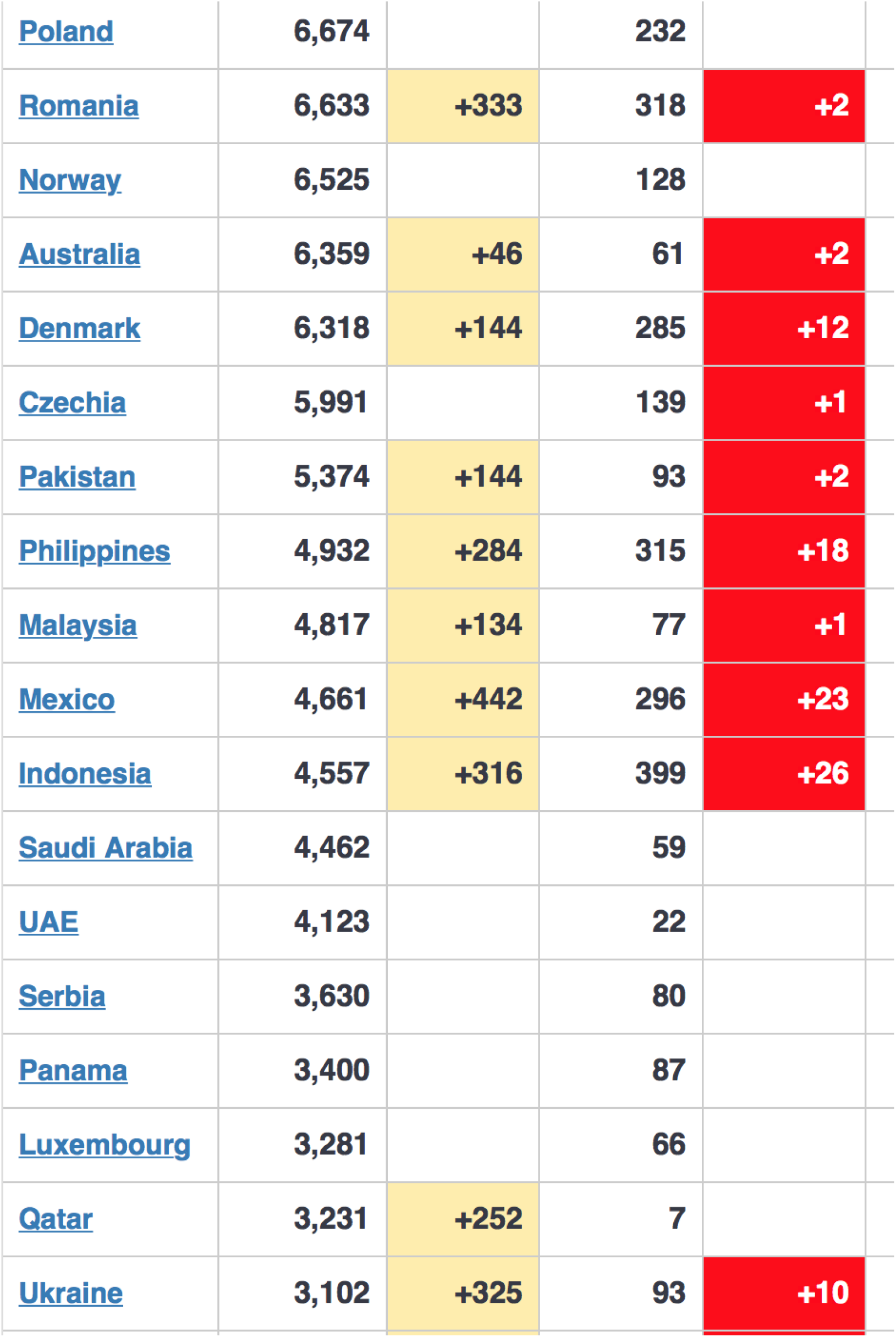
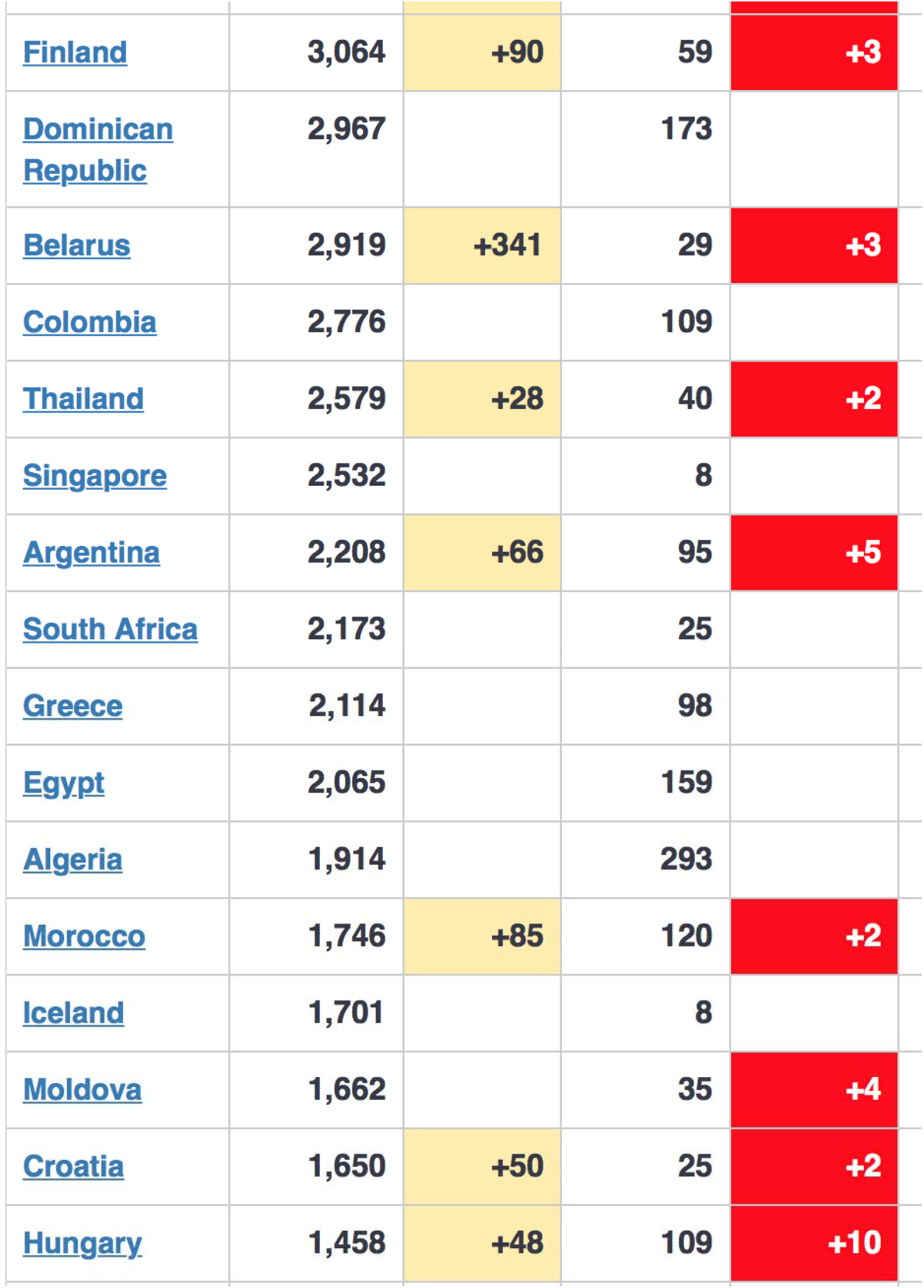
**list of the 60 countries with the most Covid 19 total cases on 13^th^ April 2020 (see note 3 of this study) from *https://www.worldometers.info/coronavirus/#countries***

## Footnotes

1 Cortegiani A, Ingoglia G, Ippolito M, Giarratano A, Einav S, *A systematic review on the efficacy and safety of chloroquine for the treatment of COVID-19*. Journal of Critical Care, 10 march 2020, available online: *https://www.ncbi.nlm.nih.gov/pubmed/32173110?fbclid=IwAR3inu2KMU9p8a1z__S3ucf2WWeFrXKzazXFcIkthX6TcOakbqseDzgeB0c*

2 Richardson Valerie, *Hydroxychloroquine rated ‘most effective therapy’ by doctors for coronavirus: Global survey*. The Washington Times, 2 April 2020. Available online: *https://www.washingtontimes.com/news/2020/apr/2/hydroxychloroquine-rated-most-effectivetherapydo/?fbclid=IwAR2e0CFgalMskxIYyAS2VFBmFGQeFyEyT3AtiASUT6O2FB9Xo5J3zOHKbcQ*

3 List available at the 13th of April (see detailed list in annex): *https://www.worldometers.info/coronavirus/#countries*

4 Hyndman R. *et al*., *Forecasting Functions for Time Series and Linear Models*, 2020. https://cran.r-project.org/web/packages/forecast/forecast.pdf

5 Box, George; Jenkins, Gwilym (1970). *Time Series Analysis: Forecasting and Control*. San Francisco: Holden-Day, 1970.

6 Gujarati Damodar N., *Econométrie*, Traduction de la quatrième édition américaine par Bernard Bernier, Collection Ouvertures économiques, éditions de Boeck, 2012.

7 Choudhary Ishan, *Forecasting COVID-19 cases in India: How many cases are going to get detected by 7th April 2020 in India?* Medium, 29 march 2020, link: *https://towardsdatascience.com/forecasting-covid-19-cases-in-india-c1c410cfc730*

8 Lardic S. et Mignon V.: *Econométrie des séries temporelles macroéconomiques et financières*, Economica, 2002.

